# Phenotyping using Structured Claims And Linked Electronic health records (PhenoSCALE): A semi-automated pipeline with an example of acute kidney injury

**DOI:** 10.1101/2025.10.23.25338672

**Authors:** Richeek Pradhan, Joyce Lii, Shirley V. Wang, Robert Ball, Rishi J. Desai

## Abstract

**Objectives:** Probabilistic phenotyping of health events has focused on unstructured or laboratory-based electronic health records (EHR), though drug surveillance still mostly relies on claims databases. Using acute kidney injury (AKI) as a case-example, we aimed to develop a semi-automated probabilistic phenotyping workflow using claims data.

**Materials and methods:** We defined highly sensitive “bronze” AKI events using ICD-10 codes and more specific “silver” events through two or more ICD-10 codes within 10 days of the bronze event. Data-driven feature selection identified co-occurring claims and the least absolute shrinkage and selection operator (LASSO) model predicting the silver labels was used to reduce the feature space and develop the final algorithms. These models were then validated against creatinine-based “gold” events derived from linked EHRs in a 20% hold out testing sample and their performance reported using area under the receiver operating characteristics curve (AUROC) and precision-recall curve (AUPRC).

**Results:** A total of 2144 features were identified based on co-occurrence with the AKI ICD codes. LASSO identified a set of 36 candidate features as most predictive of the silver labels, of which 7 were manually removed. The final phenotyping model with 29 claims-based features had an AUROC of 0.92 in the testing sample, compared to AUROC of 0.77 for a rule-based approach requiring 2 ICD codes for AKI. The AUPRC of the phenotyping model was 0.52.

**Discussion:** Phenotyping algorithm developed using our semi-automated workflow outperformed rule-based approach for identifying AKI.

**Conclusion:** This workflow may hold promise for broader application in phenotyping large-scale health data.

**Lay Summary:** Understanding whether a patient has experienced a medical event like acute kidney injury (AKI) is not always straightforward when using health insurance claims data. Traditionally, researchers use simple rules—such as the presence of diagnostic codes—to identify such events, but these methods can miss cases or include incorrect ones. In this study, we developed a new semi-automated approach to improve the identification of AKI using only claims data. We first labeled potential AKI events using diagnostic codes and then used a statistical method to select other claims records that frequently occurred around these events. These patterns were used to build a prediction model, which we tested against lab-confirmed cases from linked medical records. Our model correctly identified AKI events with high accuracy and performed significantly better than the traditional rule-based method. This approach could help researchers and public health agencies more accurately identify health events in large datasets, especially when lab results or detailed clinical notes are not available.

## Background and Significance

Phenotyping health events of interest from insurance claims or electronic health records data constitutes a key step in monitoring drug safety using healthcare databases. Accurately identified health events are essential in constructing cohorts, adjusting for covariates, and crucially, identifying safety outcomes for drug safety research and therefore, has been a focal point of attention. For example, a major project in the initial phase of the United States Food and Drug Administration (FDA)’s Sentinel initiative aiming to build a nationwide infrastructure for drug safety involved 31 academic centers and private organizations to explore the phenotype validity of 20 safety outcomes over two years.^1^ Traditionally, phenotyping activities have relied on manually developed ‘rule-based’ algorithms consisting primarily of diagnosis codes. Recently, however, there has been an increasing interest in computable phenotyping approaches to probabilistically define health events of interest using information from sources besides diagnosis codes, such as procedure codes and medications recorded as structured fields as well as features extracted from natural language processing (NLP) of free-text notes.^2^ Typically, development of probabilistic phenotyping algorithm is a time and resource intensive task which requires investigators to identify and curate clinically meaningful features based on expert knowledge.

Leveraging data-driven feature selection could make the process of phenotyping algorithm development more efficient and scalable. Approaches such as PheNorm, that are based on NLP from free text notes, have shown promise in achieving this goal.^3 4^ However, Sentinel and other surveillance networks worldwide^5^ rely heavily on structured data derived primarily from insurance claims.^6 7^ Therefore, developing a phenotyping workflow that focuses on structured data could have important practical implications. To that end, we aimed to evaluate a semi-automated process for development of phenotyping algorithms that identifies structured features in a data-driven way. As our example phenotype, we selected acute kidney injury (AKI). AKI has been identified as an important outcome for drug safety studies by both the FDA^8^ and European Medicine Agency.^9^ Prior validation studies have demonstrated that expert-curated ICD10 codes for AKI alone or in combinations with dialysis procedure codes have suboptimal performance with respect to sensitivity (median sensitivity in a systematic review of 11 validation studies being 29% and range, 15%-81%) and positive predicted value (PPV) (median, 67% and range, 15%-96%). ^10–14^ While approaches requiring dialysis as a part of the definition in prior studies generally have higher PPVs, they tend to identify only a small proportion of overall cases. More recently, a study that used the currently internationally accepted Kidney Disease: Improving Global Outcomes (KDIGO) criteria to validate AKI in electronic health records, a claims-based algorithm had a sensitivity of 52%, a positive predictive value of 39%, and an area under curve of 0.713. Therefore, we evaluated whether a scalable phenotyping approach using data-driven feature selection could be helpful in more accurately phenotyping this challenging health condition of interest from routinely collected healthcare data.

## Materials and Methods

### Setting

For this demonstration project, we used data from the Mass General Brigham (MGB) site of the FDA Sentinel Real World Evidence Data Enterprise (RWE-DE).^15^ Briefly, comprising of two major teaching hospitals, and a network of specialty and community hospitals as well as specialty practices, urgent care centers, and outpatient clinics, the MGB health system is the largest in Massachusetts. Electronic health records (EHRs) from MGB system between 2000-2020 are linked to claims from fee-for-service Medicare (2007-2020) and Medicaid (2000-2018) and have been routinely used for method development work for the FDA Sentinel Initiative. Raw claims from Medicare parts A, B, and D were converted to the Sentinel Common Data Model and required to pass strict quality assurance standards according to our routine process.^16 17^ For this study, we restricted to patients with linked EHR and Medicare claims because of recency of the claims data. This project did not require Institutional Review Board oversight as it was classified as a public health surveillance activity by the FDA and received exemption from Mass General Brigham IRB on their independent review.

### Workflow overview

We implemented a process involving automation at various steps with the ultimate objective of achieving scalability of phenotyping activities in Sentinel. For the selected phenotype of AKI, step 1 involved defining working “bronze” and “silver” events for efficient initial identification of the phenotype (**Figure 1**). “Bronze” events were intended to be highly sensitive e.g. any ICD-10 code for AKI, while “silver” events were more specific e.g. 2 or more ICD-10 codes for the phenotypes of interest within 10 days of the bronze index date. We also identified “gold” events using laboratory criteria defining AKI. Step 2 involved data-driven identification of frequently co-occurring codes (diagnoses, prescriptions, and procedures) with the “bronze” event defining criteria from structured data. Notably, this step was critical for the scalability of phenotyping as it minimized the traditionally time- and resource-consuming step of manual expert-led feature identification. Step 3 involved fitting machine learning models capable of feature selection such as penalized regression using the features identified in Step 2 as independent and “silver” events as the dependent variable to identify a smaller number of most predictive features and develop a phenotyping algorithm. Finally, in Step 4, the developed phenotyping algorithm was internally validated within a 20% hold-out set using “gold” events as the dependent and most predictive features from step 3 as independent variables of interest.

**Figure 1.** Workflow for PhenoSCALE using claims-electronic health record (EHR) linked data **\*** Gold labels are derived using serum creatinine results based on KDIGO definition of acute kidney injury. For other phenotypes, this could involve manual review of charts.

#### Step 1: Initial phenotype identification

##### a. Base cohort (Figure 2A)

Patients receiving care in the MGB healthcare system between October 2015 and December 2020 and enrolled in Medicare for at least one month were eligible for the study. The base cohort included individuals who had reached their 18^th^ birthday by October 1, 2015, with the cohort entry date being the latest of the two dates, 18^th^ birthday and October 1, 2015. Patients were followed up until death, the end of data in the system, or December 31, 2020, whichever occurred earliest.

**Figure 2.** Phenotyping and feature accrual. 2A. Step 2: Initial phenotype identification: Constructing the base cohort; 2B. Step 2: Initial phenotype identification: Identifying bronze events; 2C. Step 2: Initial phenotype identification: Identifying silver events; 2D. Step 2: Initial phenotype identification: Silver events identified; 2E. Step 2: Initial phenotype identification: Risk set sampling for control events; 2F. Step 2: Initial phenotype identification: Full risk set matched base cohort; 2G. Step 2: Initial phenotype identification: Scanning for creatinine value within feature accrual period; 2H. Step 2: Initial phenotype identification: Identifying those meeting AKI criteria from among those who had at least one creatinine record; 2I. Step 2: Initial phenotype identification: Demarcating creatinine-validated gold events and control events; 2J. Step 2: Feature accrual: Identification of co-occurring codes in feature assessment window; 2K. Step 2: Feature accrual: Identification of co-occurring codes.

##### b. Identification of bronze events (Figure 2B)

Patients from the base cohort were further evaluated for an ICD-10 code for AKI. A set of ICD-10 codes (N17.0 (acute renal failure with tubular necrosis), N17.1 (acute renal failure with acute cortical necrosis), N17.2 (acute renal failure with medullary necrosis), N17.8 (Other acute renal failure), N17.9 (acute renal failure, unspecified)) with the highest sensitivity, based on published validated studies,^18^ was used as the bronze event to identify all potential AKI episodes, that is, bronze events. The first day of the encounter during which the ICD code was recorded was considered the bronze index date to identify the silver events and operationalize the feature assessment window.

##### c. Defining silver label events (Figure 2C,D)

Patients in whom bronze events identified were further evaluated to determine if they qualified as having silver events. The silver events are likely events with higher specificity but lower sensitivity. We identified a subset of bronze events who had two traditional AKI ICD-10 codes (N17.0, N17.1, N17.2, N17.8, N17.9) within 10 days of the bronze index date as silver events for use model development for dimension reduction (Step 3).

##### d. Sampling control events (Figure 2E, F)

To select controls, we used a risk set sampling approach from the base cohort. For each patient with a newly recorded AKI diagnosis (bronze event), we identified potential controls from the base cohort free from AKI and had follow-up time equal to or longer than the bronze event. Up to 10 controls were randomly sampled for each bronze event from this risk set and assigned an index date so that the control and their corresponding bronze event had the same base cohort follow-up. By design, a patient could serve as a control for multiple bronze events and could later become a bronze event. The rationale behind selecting control events was to facilitate feature identification in Step 2 by contrasting prevalence of co-occurring codes between bronze events and control events.

###### d. Defining gold events (Figure 2G,H,I)

To define gold events, we used the creatinine values modified from the KDIGO criteria (**Supplementary methods 1**). First, we selected patients with bronze events and control events who had at least one creatinine value within −10 days and up to +10 days of the bronze index date, to ensure that everyone within this selected set had similar opportunity to be evaluated for the KDIGO criteria. Within this set, we identified those meeting the modified KDIGO criteria as having creatinine-validated gold events and those not meeting these criteria during their feature assessment window as creatinine-validated control events. Gold events were used only for validation in Step 4.

#### Step 2. Feature accrual (Figure 2J,K)

To find additional features beyond the AKI codes that might be highly associated with AKI events, Step 2 entailed identification of structured codes, including international classification of diseases (ICD-10, diagnosis and procedure codes), national drug code (NDC), and current procedural terminology (CPT) codes frequently co-occurring with the bronze label events within −10 days and up to +10 days of the bronze index date (feature assessment window). First, from all the codes occurring in the 20-day feature assessment window, to increase specificity by only including codes that are relevant for the ongoing AKI episode, we excluded codes which also occurred outside the feature assessment window from further consideration. We operationalized frequent co-occurrence by calculating odds ratios for all codes appearing in the feature assessment window for the bronze events and their selected control events as described above. Features with odds ratios confidence limits above the null value of 1, which would indicate greater likelihood of the code being present among bronze events compared controls, were retained. .

#### Step 3. Feature set dimension reduction and model training on silver labels

Next, we randomly separated the entire matched case control set into a 80% training set and a 20% test set. We used the 80% training set to conduct the penalized regression. We used least absolute shrinkage and selection operator (LASSO) penalized regression with silver labels as the outcomes and identified candidate features in all data dimensions as predictors. We used the SAS 9.4 HPGENSELECT with 10-fold cross validation to conduct LASSO-based feature selection. We manually reviewed the features selected by LASSO and removed the ones that are unlikely to be clinically relevant. We used the candidate features retained after this step along with AKI ICD10 codes to create the final phenotyping algorithm. The modeling used diagnostic and procedural codes to predict and classify silver events (positive cases). We calculated a series of linear predictors by applying LASSO-determined weights to each feature, representing the strength and direction of their association with the outcome. Next, the model computed probabilities for each classification (silver = 0 or 1) using logistic regression. It calculated these probabilities by applying the logistic function to the sum of the weighted predictors. Based on the higher probability, the model assigned a final classification to each case.

#### Step 4. Validation in a hold-out sample

The final phenotyping algorithm was validated in the 20% test set against creatinine-validated gold event and creatinine-validated gold non-event labels as described in Step 1d. Area under the receiver operating characteristics curve (AUROC) and precision-recall curve (AUPRC)^19^ were reported as measures of model performance. As comparison, we also reported AUROC for the rule-based approach of identifying AKI which required 2 ICD10 codes, as in our silver label definition, with the index date the first of the two events.

## Results

### Initial phenotype identification

The base cohort comprised of 493,993 patients with a total follow-up of 1,707,764 person-years. Among them, a total of 81,032 bronze events were recorded, yielding a bronze event incidence rate of 4.81 (95% confidence interval (CI) 4.77-4.84) per 100 person years. Of the individuals who had a bronze event, 42,590 met the criteria of having an additional AKI code, yielding a silver event incidence rate of 2.51 (95% confidence interval 2.48-2.53) per 100 person years. Using risk-set sampling, a total of 75,127 individuals with bronze events were matched to 751,257 control events during the risk set. Within this matched set of bronze events and their controls, 94,983 had a creatinine value during the feature accrual period, and 8017 were creatinine-validated gold events, and 86,966 were creatinine-validated control events.

**Table 1** describes the baseline characteristics patients with matched bronze, silver, and gold events. Overall, those with silver events were very similar in characteristics to those with bronze events but had marginally higher prevalence of conditions that can be clinically confused with AKI. Those with gold events were marginally younger, and less likely to be female and white than those with bronze events. They also had a higher proportion of differential diagnoses such as heart failure, chronic kidney disease, and urinary tract obstruction but a lower proportion of hypovolemia. Finally, those with gold events had higher proportion of dialysis and death, though similar prevalence of hospitalization to those with bronze and silver events.

**Table 1.** Characteristics of the bronze, silver, and gold acute kidney injury (AKI) events.

|  | Variable | Matched bronze AKI events | Matched bronze controls | Matched silver AKI events | Matched silver controls | Matched gold AKI events | Matched gold controls |
| --- | --- | --- | --- | --- | --- | --- | --- |
| N | N | 75,127 | 7,51,257 | 40,775 | 4,07,742 | 8017 | 86,966 |
| Demographic | Age,<sup>1</sup> mean (standard deviation) | 74.8 (14.2) | 64.1 (19.9) | 75.7 (13.6) | 64.3 (19.8) | 72.5 (14.9) | 69.1 (16.2) |
|  | Female,<sup>2</sup> n (%) | 37,671 (50.1) | 4,67,965 (62.3) | 20,499 (50.3) | 2,53,969 (62.3) | 3678 (45.9) | 49,323 (56.7) |
|  | White,<sup>2</sup> n (%) | 64,370 (85.7) | 5,62,453 (74.9) | 35,574 (87.2) | 3,06,010 (75.0) | 6390 (79.7) | 68,518 (78.8) |
|  | Black,<sup>2</sup> n (%) | 4410 (5.9) | 43,707 (5.8) | 2206 (5.4) | 23,571 (5.8) | 716 (8.9) | 5204 (6.0) |
| Presence of relevant comorbidities<sup>3</sup> | Diabetes, n (%) | 33,052 (44.0) | 1,49,496 (19.9) | 18,478 (45.3) | 81,438 (20.0) | 3754 (46.8) | 25,235 (29.0) |
|  | Diabetic nephropathy, n (%) | 14,447 (19.2) | 28,821 (3.8) | 8521 (20.9) | 15,891 (3.9) | 2076 (25.9) | 6919 (8.0) |
|  | Hypertension, n (%) | 67,893 (90.4) | 4,13,744 (55.1) | 37,395 (91.7) | 2,26,618 (55.6) | 7310 (91.2) | 61,863 (71.1) |
|  | Hypertensive nephropathy, n (%) | 31,006 (41.3) | 39,709 (5.3) | 18,315 (44.9) | 21,835 (5.4) | 4334 (54.1) | 11,126 (12.8) |
| Presence of alternative diagnoses<sup>4</sup> | Heart failure, n (%) | 26,326 (35.0) | 11733 (1.6) | 15,624 (38.3) | 6372 (1.6) | 3451 (43.0) | 6299 (7.2) |
|  | Chronic kidney disease, n (%) | 39,638 (52.8) | 75,982 (10.1) | 22,767 (55.8) | 41,713 (10.2) | 5057 (63.1) | 17441 (20.1) |
|  | Urinary calculi, n (%) | 4423 (5.9) | 3449 (0.5) | 2755 (6.8) | 1873 (0.5) | 444 (5.5) | 1350 (1.6) |
|  | Hypovolemia, n (%) | 22,410 (29.8) | 2508 (0.3) | 14,543 (35.7) | 1361 (0.3) | 1901 (23.7) | 4663 (5.4) |
|  | Urinary tract infection, n (%) | 17,967 (23.9) | 12,259 (1.6) | 11,273 (27.6) | 6628 (1.6) | 1725 (21.5) | 4720 (5.4) |
|  | Urinary tract obstruction, n (%) | 3358 (4.5) | 1041 (0.1) | 2350 (5.8) | 580 (0.1) | 393 (4.9) | 717 (0.8) |
| Severity of the event | Dialysis,<sup>4</sup> n (%) | 2119 (2.8) | 2222 (0.3) | 1343 (3.3) | 1202 (0.3) | 733 (9.1) | 302 (0.3) |
|  | Hospitalization,<sup>4</sup> n (%) | 61,291(81.6) | 12,200 (1.6) | 36,523 (89.6) | 6754 (1.7) | 7281 (90.8) | 14,703 (16.9) |
|  | Death within 30 days,<sup>5</sup> n (%) | 6148 (8.2) | 1245 (0.2) | 3644 (8.9) | 667 (0.2) | 1268 (15.8) | 1303 (1.5) |

<sup>1</sup> Measured on index date
<sup>2</sup> Measured on index date based on recording by CMS
<sup>3</sup> Measured on or a month before base cohort entry date to the end of feature accrual period in claims
<sup>4</sup> Measured during feature accrual period in claims
<sup>5</sup> Measured on or within 30 days from index date

### Feature accrual

Within the feature assessment window of the cases, we found a total of 20,269 ICD-10 diagnosis codes, 6,508 ICD-10 procedure codes, 15,007 NDC codes, and 5,282 CPT codes. Of these 2,025 ICD-10 diagnosis codes, 37 ICD-10 procedure codes, 6,081 NDC codes, and 284 CPT codes were excluded as they had appeared both inside but also before the feature assessment window among the cases, leaving a total of 38,634 codes. Among these codes, 1,231 ICD-10 diagnosis codes, 34 ICD-10 procedure codes, 356 NDC codes, and 523 CPT codes yielded odds ratios with confidence intervals above the null when compared to their co-occurrences among control events.

### Feature set dimension reduction and model training on silver labels

At this stage, we separated the data into 80% training set and 20% testing set. Within the 80% training data with 32,497 silver events, we conducted LASSO regression with the silver events as the outcome variable and 2144 features identified in the feature accrual step as predictors. LASSO selected 36 features (26 ICD-10 diagnosis codes, 2 ICD-10 procedure codes, and 8 CPT codes) out of the 2144 total candidates as most predictive of silver events. After manual review of the retained codes, we further excluded 7 administrative codes that were unlikely to be clinically relevant or specific to AKI (ICD-10 Z66: Do Not Resuscitate status, CPT 99221-99223: initial inpatient or observation care, 99233: subsequent inpatient or observation care, 99239: hospital inpatient or observation discharge services, 99291: critical care services) to reduce the feature space to 29 distinct codes (**Table 2**).

**Table 2.** International Classification of Diseases (ICD) and Current Procedural Terminology (CPT) codes used in the final phenotyping algorithm.

| **Code type** | **Codes** | **Description** |
| --- | --- | --- |
| ICD-10 diagnosis codes | A41.9 | Sepsis, unspecified organism |
|  | D72.829 | Elevated white blood cell count, unspecified |
|  | E86.0 | Dehydration |
|  | E86.1 | Hypovolemia |
|  | E87.1 | Hypo-osmolality and hyponatremia |
|  | E87.2 | Acidosis |
|  | E87.5 | Hyperkalemia |
|  | E87.6 | Hypokalemia |
|  | G93.41 | Metabolic encephalopathy |
|  | I12.9 | Hypertensive chronic kidney disease with stage 1 through stage 4 chronic kidney disease, or unspecified chronic kidney disease |
|  | I95.9 | Hypotension, unspecified |
|  | J90 | Pleural effusion, not elsewhere classified |
|  | J96.01 | Acute respiratory failure with hypoxia |
|  | J98.11 | Atelectasis |
|  | N18.9 | Chronic kidney disease, unspecified |
|  | N19 | Unspecified kidney failure |
|  | N28.9 | Disorder of kidney and ureter, unspecified |
|  | R06.02 | Shortness of breath |
|  | R07.9 | Chest pain, unspecified |
|  | R09.02 | Hypoxemia |
|  | R41.82 | Altered mental status, unspecified |
|  | R50.9 | Fever, unspecified |
|  | R53.1 | Weakness |
|  | R79.89 | Other specified abnormal findings of blood chemistry |
|  | Z45.2 | Encounter for adjustment and management of vascular access device |
| ICD-10 procedure codes | 02HV33Z | Insertion of infusion device into superior vena cava, percutaneous approach |
|  | 30233N1 | Transfusion of nonautologous red blood cells into peripheral vein, percutaneous approach |
| CPT codes | 74176 | Computed tomography, abdomen and pelvis; without contrast material |
|  | 76770 | Ultrasound, retroperitoneal (eg, renal, aorta, nodes), real time with image documentation; complete |

### Validation in a hold-out sample

Finally, within the 20% test set, a total of 20,208 observations had at least one creatinine measurement in their feature assessment window. Of these, 1624 events (8%) were labeled as creatinine-validated gold events and 18,584 (92%) as creatinine-validated control events. In this validation sample of creatinine-validated gold events and control events, the final phenotyping model, had an AUROC of 0.92 (95% CI 0.92-0.93), compared to AUROC of 0.77 (0.76-0.78) for a rule-based approach requiring 2 ICD codes for AKI (**Figure 3**). The AUPRC of the phenotyping model was 0.52 (**Figure 4**).

**Figure 3.** The receiver operating characteristics curve in the testing data using laboratory-based definition as the gold standard.

**Figure 4.** Precision-recall curve for the phenotyping model in the testing data using laboratory-based definition as the gold standard.

## Discussion

Using AKI as a case-example, we outlined PhenoSCALE, a novel workflow to select structured features from information recorded in claims data to build computable phenotyping algorithms. Manual review confirmed that most of the features selected by our data-driven approach in PhenoSCALE were clinically plausible and relevant for AKI. Compared to a traditional rule-based approach of using ICD codes alone, the computable phenotyping algorithm demonstrated substantially higher AUROC.

Our work is an extension of the approaches currently available in the literature for semi-automated phenotyping, most notably PheNorm, which also uses noisy silver labels for model training.^3^ In contrast to PheNorm, which introduces automation in selection of NLP based features from free-text notes using an automated feature engineering pipeline, we focus on automating selection of features based on information recorded in insurance claims data as structured fields. Specifically, after identifying all co-occurring codes with the bronze labels, we exclude features that are recorded in the history of the patients, thus reducing the possibility of chronic risk factors emerging as candidate predictors, thus increasing efficiency as well as specificity. Next, we restrict to features with confidence limits for the odds ratio contrasting frequency of these codes between bronze events and non-events derived using a risk set sampling approach above the null value of 1.0, which ensures inclusion of features that co-occur frequently enough to materially add to the predictive ability of the final algorithm. Finally, the use of a risk set sampling approach to sieve out codes has several advantages. For example, this approach selects controls from individuals may develop the condition of interest later, thus avoiding selection of fundamentally healthier controls.^20 21^ Moreover, this also allows for matching on additional factors, such as demographic variables, treatment characteristics, or differential diagnoses, which can help control for confounders and improve the precision of the phenotyping process.

It is noteworthy that our approach narrowed down the set of candidate features from a total of 47,066 co-occurring codes to 36, eliminating 99.92% of the codes, entirely automatically. Automation in feature selection and retention addresses one of the most important bottlenecks, which is labor-intensive manual feature curation, common to traditional phenotyping algorithm development workflows. It could also potentially minimize variability introduced by operator dependent manual processes and ensures consistent and scalable application across large datasets. However, this also introduces potential variability in investigator choices in terms of when to capture co-occurring codes, and how to feed in the inputs for LASSO to choose from (amount of pre-processing, e.g. generic names versus medical classes), which should be well-documented while implementing such a workflow.

The codes that were shortlisted for the final probabilistic phenotyping algorithm for AKI included signs and symptoms of AKI, metabolic changes associated with AKI, signs and symptoms or diagnosis of acute causes of AKI, chronic kidney disease codes, and investigations and common treatments associated with AKI. Overall, the selected codes had clinical face-validity, which lends credibility to our approach of entirely data-driven feature selection. However, notably some obvious signs and symptoms of AKI such as anuria and oliguria did not emerge as candidate features. A possible explanation is that such factors are not coded as frequently or as reliably in claims data and hence are unlikely to add value for probabilistic phenotyping. While the present code list performed superior with respect to ROC compared to AKI codes, further refinement of the selection process may be achieved if the risk set controls were selected among those who had hospitalization or other clinical encounters (to further weed out encounter associated nonspecific codes) more comparable in terms of age, sex and calendar time, by including these variables in the regression model (so as to make the selected codes more comparable), without compromising the automaticity of the feature accrual process. While in this proof-of-concept pipeline feasibility study, we did not include these steps, they should be considered in future analyses.

While previous studies have used chart review to validate AKI, we compared our study results with Zhang et al. (2022), which is the only previous study that also used KDIGO criteria as the gold standard and is based on ICD10 codes (as opposed to ICD9) similar to our study. ^10^ Zhang et al. examined various rule-based algorithms based on one or more AKI codes, AKI-related procedure codes, and unstructured mentions of AKI-related terms in clinical notes (ROC ranging between 0.710-0.743). A key similarity is observed in the ROC performance for ICD-based codes in identifying AKI; both studies yield comparable ROC values, indicating similar accuracy of ICD codes for initial AKI identification in the datasets. However, our model did not select features related to renal replacement therapy, including dialysis. This may be due to the predominance of AKI cases managed without dialysis, suggesting that incorporating these codes may not meaningfully improve sensitivity in AKI identification. Furthermore, our study demonstrates the strength of our probabilistic algorithm, which outperforms the ICD-based algorithm with respect to the ROC, indicating that our method more accurately classifies AKI cases and captures a broader range of cases.

Some limitations of our work merit discussion. First, the proposed workflow is unlikely to be applicable to certain phenotypes that are not well-defined by existing codes or patterns that can be used as reasonable silver labels or when no gold labels are available. Second, while efficient and timely, the proposed semi-automated workflow may also not be sufficient for instances where more clinical expertise is required in selecting or prioritizing certain features over others for the purposes of sub-phenotyping events of interest. For instance, the general features picked up by our workflow for AKI may not be ideal if the goal was to differentiate between varying etiologies of AKI such as drug-induced versus resulting from other clinical complexities such as sepsis. Third, we conducted the validation using data from the same healthcare system, so we cannot comment on the transportability of the algorithm in a truly external validation sample from a separate site. Fourth, given the internationally accepted criteria to define AKI are based on laboratory values, we used these criteria to identify the gold cases. However, gold cases may also be identified using chart review, particularly in case of clinically diagnosed health events of interest. Whether this pipeline approach is effective in feature accrual in such cases needs to be examined in future projects. Fourth, since serum creatinine is a diagnostic criterion for AKI, it would typically precede the AKI diagnosis in the usual clinical workflow. However, we acknowledge that this may not always be the case in practice. For instance, a physician might initially assign an AKI diagnosis based on urine output or other clinical signs and symptoms and then confirm it later with laboratory results. Similarly, administrative processes could result in laboratory measurements being recorded after the diagnostic code is assigned. To address this, we required serum creatinine measurements within a 10-day window on either side of the recorded diagnosis to validate the AKI diagnosis. However, there may have been rare instances where creatinine was not recorded—either due to missed testing or database artifacts. Finally, this approach relies on dense information content during the feature accrual period and therefore, certain patients, for instance those who die soon after recording of the ‘bronze’ code, could preferentially have less opportunity to be classified as having an event because they have less follow-up time for collection of additional codes.

## Conclusion

In summary, our automated probabilistic phenotyping approach offered strong face-validity and higher accuracy compared to traditional rule-based approach. This approach holds promise for improving the identification of clinical phenotypes using structured features from claims data in large-scale database studies.

## COMPETING INTEREST STATEMENT

Dr. Desai has served as Principal Investigator on research grants from Vertex, Novartis, and Bristol Myers Squibb to Brigham and Women’s Hospital on unrelated projects. Dr. Ball is an author on US Patent 9,075,796, ‘Text mining for large medical text datasets and corresponding medical text classification using informative feature selection’. At present, this patent is not licensed and does not generate royalties.

## CONTRIBUTIONS

All authors conceived and designed the study. RD acquired the data. RP, JL, and RD did the statistical analyses. All authors analysed and interpreted the data. RP and RD wrote the manuscript, and all authors critically revised it. All authors approved the final version of the manuscript and agree to be accountable for the accuracy of the work. RD supervised the study and is the guarantor.

## FUNDING

This project was funded by Task Order 75F40122F19006 under Master Agreement 75F40119D10037 from the US Food and Drug Administration (FDA).

## DATA AVAILABILITY STATEMENT

The data underlying this article cannot be shared publicly due to institutional policies that protect the privacy of individuals whose data was used in the study.

## DISCLAIMER

FDA coauthors reviewed the study protocol, statistical analysis plan, and the manuscript for scientific accuracy and clarity of presentation. Representatives of the FDA reviewed a draft of the manuscript for presence of confidential information and accuracy regarding statement of any FDA policy. The views expressed are those of the authors and not necessarily those of the US FDA.

## Notes

### Author Declarations

This project did not require Institutional Review Board (IRB) oversight as it was classified as a public health surveillance activity by the FDA and received exemption from Mass General Brigham IRB on their independent review.

