## Supplementary methods 1 for "Phenotyping using Structured Claims And Linked Electronic health records (PhenoSCALE): A semi-automated pipeline with an example of acute kidney injury"

**Supplementary methods 1. Defining creatinine-validated gold events**

**Kidney Disease: Improving Global Outcomes (KDIGO) criteria for acute kidney injury**

- Increase in serum creatinine by 0.3 mg/dL or more (26.5 μmol/L or more) within 48 hours.
- Increase in serum creatinine to 1.5 times or more than the baseline of the prior 7 days.
- Urine volume less than 0.5 mL/kg/h for at least 6 hours.

**Modifications to identify acute kidney injury in the database based on creatinine values**

Examine only among bronze events and control events, those who have at least one creatinine value. Within them, individuals can be flagged as gold events if:

1.	They have within the feature accrual period (– 10 to <= +10 days of the bronze index date) a creatinine value ≥1.5 times the upper limit of the normal range of the lab report

OR

2.	They have within the feature accrual period two creatinine values within 2 days of each other where the difference between the earlier and later values is ≥0.3 mg/dL (≥26.5 micromol/L).
